# From Fragmented to Functional: Improving Rehabilitation Data in Georgia’s Health Information Systems for Better Decision-Making

**DOI:** 10.1101/2025.06.02.25328247

**Authors:** Nino Kotrikadze, Akaki Zoidze, George Gotsadze

## Abstract

**Introduction:** Integrating rehabilitation data into health information systems (HIS) is critical for effective healthcare decision-making, especially in low- and middle-income countries (LMICs) like Georgia. Despite legal obligations under the Convention on the Rights of Persons with Disabilities (CRPD) and the Georgia Rehabilitation Service Development Strategy (2023–2027), rehabilitation data in Georgia remains fragmented. This study aims to assess the adequacy of the HIS for rehabilitation in Georgia to support the planning, monitoring, and expansion of rehabilitation service developments in the country.

**Methods:** The cross-sectional study design combined a documentary review with qualitative interviews. The documentary review examined legislative/regulatory documents from official bodies. Qualitative data were collected through 24 semi-structured interviews with stakeholders representing six rehabilitation facilities (four providing services under the Universal Health Care Program (UHCP) with government funding and two on a fee-for-service basis) and two policymakers. NVivo 12 was used to analyze interviews through the first deductive (Health Metrics Network framework) followed by an inductive coding approach.

**Results:** Findings reveal that rehabilitation data in Georgia’s HIS are inconsistent, lacking standardized definitions or reporting practices. Differences were identified in data collection practices across facilities. The UHCP facilities employ the functional assessment tool but reporting inconsistencies persist. Multiple recording modules lack standardization, with variations in the volume and type of data reported. Key challenges include the absence of unified coding systems, incomplete data capture, and reliance on unstructured formats, hindering data analysis and integration.

**Conclusion:** Although Georgia has made progress by integrating rehabilitation services into the UHCP, gaps in HIS still limit effective decision-making. Addressing these gaps requires a standardized data collection framework emphasizing programmatic and functional outcomes, patient-centered measures, and the comprehensive use of existing electronic systems. Strengthening HIS via coordinated stakeholder efforts and streamlined data reporting will improve rehabilitation services and serve as a model for other LMICs.

## Introduction

As global populations age and the incidence of chronic disease increases, many individuals face functional limitations that impede daily activities and social participation (1). Between 1990 and 2019, the number of years people lived with disabilities globally increased by 63%, indicating a significant rise in the need for rehabilitation services. By 2021, 2.6 billion people—approximately one-third of the world population—required rehabilitation due to disease or injury, further intensifying the demand for healthcare services (2–4).

The World Health Organization (WHO) launched the Rehabilitation 2030 initiative in 2017 in response to the growing challenge. This global “call to action” urges countries to prioritize rehabilitation as a core component of health systems, aiming to improve access and the quality of rehabilitation services worldwide (5). However, fewer than 50% of individuals receive the necessary rehabilitation services today (2) because health systems face several challenges in scaling up services, particularly in low- and middle-income countries (LMICs). Lack of accurate population data and inadequate information systems not capturing disability-related information (only 31% of datasets from 133 LMICs included relevant facts, and just 15% used internationally standardized functional difficulty questions) (6) imposes significant limitations on planning for service creation/expansion. Moreover, access to rehabilitation services in LMICs remains limited, with shortages in rehabilitation service providers, assistive devices, and effective therapies. These issues are compounded by a global shortage of rehabilitation professionals, especially in rural areas (7) and inadequate health financing schemes, leading to high out-of-pocket costs for those in need of rehabilitation (8).

Without adequate health information systems (HIS), planning for rehabilitation service development and integration in the health system and its eventual expansion to meet the population’s needs would not be possible. To help countries, the WHO developed and recommended a set of monitoring and evaluation indicators (9) to be generated through national HIS to plan and monitor rehabilitation service development and delivery expansions. An accurate and comprehensive HIS has to provide information about the health status/needs of the population and system performance data, supporting evidence-based decision-making for rehabilitation service development (10). Therefore, our study aims to assess the adequacy of the HIS for rehabilitation in Georgia to support the planning, monitoring, and expansion of rehabilitation services in the country.

In Georgia, rehabilitation services have traditionally been tied to disability, an inheritance from the Soviet past. These services were funded through social care programs/budgets with little or no relevance to healthcare. After independence in 1991 and in light of significant economic decline, when Georgia’s Gross Domestic Product (GDP) fell by 78% during 1990-1995 (11), Georgia retained some public funding primarily serving children with disabilities, while adults received minimal support in the form of assistive technologies (12) and faced significant out-of-pocket costs and were exposed to financial access barriers when seeking rehabilitative services. In 2013, the Government of Georgia introduced the Universal Healthcare Program (UHCP), which expanded public coverage to almost the entire population, many of whom were not entitled to state support before this program. The UHCP aimed to provide more equitable access to healthcare services across socioeconomic groups but not cover rehabilitative services. The depth of coverage under the UHCP for lower-income households is higher, with more extensive benefits.

In particular, pensioners, children aged 0–5, and families living below the poverty line are prioritized for comprehensive medical coverage. These groups gained access to a wide range of essential healthcare services, including basic primary care, diagnostic services, emergency outpatient and inpatient care, elective surgeries, oncological treatments, and maternal health services like obstetric care (13). However, rehabilitation services were initially excluded, leaving a significant coverage gap. Progress was only made in November 2022, when financing for adult rehabilitation services were integrated into the UHCP, with the first phase providing coverage for conditions such as stroke, traumatic brain and spinal cord injuries and with the plan for future expansion of the covered conditions (14).

Several important events preceded the integration of rehabilitation services into Georgia’s health system. One of the most important milestones was Georgia’s ratification of the Convention on the Rights of Persons with Disabilities (CRPD) in December 2013, demonstrating Georgia’s political commitment to improving rehabilitation services (15). After that, in 2020, the WHO, in collaboration with the Ministry of Internally Displaced Persons from the Occupied Territories, Labour, Health and Social Affairs (MoH), conducted a situational analysis of Georgia’s rehabilitation sector, revealing critical areas for immediate attention, such as poorly integrated services into the health system, an underdeveloped workforce, and a lack of comprehensive data on rehabilitation outcomes, quality, and efficiency (12). To address systemic shortcomings, the WHO helped Georgia elaborate the National Strategy for the Development of Rehabilitation Services (2023–2027), which the government approved on January 30, 2023 (16). The strategy outlines objectives for developing rehabilitation services and establishes 14 indicators to be monitored within the health system to improve data collection and ensure evidence-informed decision-making.

Thus, a robust HIS became critical as Georgia integrated rehabilitation services into the health system. Therefore, this paper describes the current information flow that must supply the indicators for the Georgia Rehabilitation Service Development Strategy (2023–2027), identifies the strengths and weaknesses of information collection, reporting and analysis and elaborates on recommendations for system enhancement that could also be relevant for other countries embarking on a similar journey.

## Methodology

Due to the limited prior research on rehabilitation HIS in Georgia, a cross-sectional, exploratory design was chosen. Primary qualitative data collection was integrated with a documentary review to achieve the study objectives. This approach allows a comprehensive view of current rules, norms and practices (17) and helps identify strengths and areas for improvement. The document review covered legislative and regulatory documents and published and grey papers, while qualitative interviews were conducted with purposefully selected key stakeholders, including policymakers and employees of the facilities involved in rehabilitation service provision, with and without government funding.

To capture a broad range of perspectives, purposive sampling targeted facilities that had recently joined Georgia’s UHCP as rehabilitation service providers in November 2022 and the providers operating without government funding. Of the six facilities registered as UHCP providers at the time, four agreed to participate in the study. Additionally, two non-UHCP facilities were included to compare data collection and reporting approaches between UHCP and non-UHCP providers. This approach offered valuable insights into data management practices.

Key informants involved in rehabilitation policy and data governance were also purposefully selected. In total, 24 interviews were conducted: 22 with healthcare facility representatives and 2 with policymakers. Facility participants included physicians, physical therapists, personnel involved in human resource management, quality managers, and statisticians, ensuring professional representation of diverse groups. All invited participants agreed to participate, and none withdrew from the study.

Data collection occurred in two geographic locations: four rehabilitation facilities in the capital city of Tbilisi and two in the regions. Of the 24 interviews, 22 were conducted face-to-face, and two were conducted virtually via Microsoft Teams to accommodate participants’ availability. Interviews took place in private settings within the facilities to ensure confidentiality and minimize distractions. Only the interviewer and participant were present during each interview.

Semi-structured interviews were used for data collection. The interview guide was developed based on the Health Metrics Network (HMN) Framework and pilot-tested with 3 participants to ensure clarity and relevance of asked questions (18). Minor adjustments were made based on pilot feedback, such as refining questions related to data reporting practices to ensure better alignment with participants’ roles. All interviews were audio-recorded after securing the participant’s written consent, and field notes were taken to capture additional context during and after each session. Ethical approval was obtained from the National Bioethics Committee of Georgia (Ethical approval (IRB # 2023-074)) housed at the National Center for Disease Control and Public Health (NCDC) before the commencement of the fieldwork. The interviews lasted between 30 and 90 minutes, depending on participant availability and the depth of discussion. Data saturation was reached after 22 interviews, with no new themes emerging in the later ones. Audio recordings were transcribed verbatim, and audio files and transcripts were securely stored on password-protected devices. Only the research team had access to the raw data, and all identifying information was removed during transcription to ensure participant confidentiality.

The transcribed interviews were analyzed using NVivo 12™. Both deductive and inductive approaches were applied. Deductive analysis followed the HMN framework, which structured the data into three main components: HIS governance/institutional roles, data sources, and data management. The inductive analysis allowed additional themes to emerge that helped elaborate/expand the deductive code tree. Coding was performed by the lead researcher, with reviews carried out by a second coder to ensure consistency and reliability. Reflexivity was maintained throughout the process. Regular meetings were held between researchers to resolve coding discrepancies and ensure consistent data interpretation. For greater robustness, the triangulation was employed across interview data and findings from the document review. The study’s findings were further validated through stakeholder dialogue aimed at reviewing the research results on rehabilitation data collection and reporting within Georgia’s HIS and deliberating on recommendations for future actions. Workshop participants included representatives from rehabilitation service providers, the WHO country office, academia, and officials from the MoH and the NHA. This approach ensured that the conclusions drawn were grounded in evidence emerging from the study and recommendations well-contextualized and resonated with the stakeholders.

## Results

### Institutions Involved in Rehabilitation HIS

This section outlines Georgia’s institutions involved with rehabilitation data management, specifying their distinct roles and responsibilities.

The National Statistics Office of Georgia (GeoStat) collects disability data through national censuses to estimate the population’s needs. In the 2014 census, GeoStat collected information on disability and functional impairment using a survey based on the Washington Group’s questions on Disability. Two key questions addressed difficulties in daily activities—such as seeing, hearing, and walking—and classified disability status into categories: none, mild, moderate, or severe, with an additional category for children with disabilities (19).

.The MoH owns and regulates a nationwide Electronic Health Records (EHR) system, which was launched in 2019. Starting from 2023, all facilities must share patient health data and financial data for cases paid by UHCP. Only cases funded by UHCP are subject to mandatory reporting (20) for rehabilitation services. The MoH also manages an electronic reporting system (minimal wage module) for inpatient facilities to document staff hours worked and wages paid. In 2023, the ministry introduced minimum wage standards for physicians and nurses working in facilities under UHCP, and this information module helps enforce compliance with new regulations (14,21).

The National Center for Disease Control and Public Health (NCDC), a leading public health institution in the country, collects data from medical facilities through routine electronic forms, such as Form No. 025, which records new cases seen by outpatient care providers, the System User’s Electronic Module (e-health users/ Form No. 079), which collects human resource data on a provider level and the Annual Healthcare Facility Report (Form No 1), which gathers healthcare facility information on available equipment, staffing, and delivered services (types and volumes) (22).

The National Health Agency (NHA), a single national service purchaser for health, funds UHCP services, including rehabilitation. NHA manages provider reimbursement through an electronic reporting system, case registration module, that collects claims data from healthcare facilities and enables online case submissions for UHCP-funded medical and rehabilitative services (23,24).

Rehabilitation service providers (public or private alike) must comply with all state regulations, report beneficiaries’ information in the system, and provide required data through NCDC reporting modules. Facilities contracted by NHA for the UHCP benefits delivery also report claims data to NHA (20). The following sections discuss detailed reporting practices observed on a facility level.

### Data Collection for Rehabilitation Services

This section focuses on institution-based data sources, including individual, service (facility records), and resource records, as depicted in Figure 1 and further elaborated in Figure 2. It does not cover population-based data sources, such as census data, civil registration, or population surveys. It explores data collection practices at the facility level and reveals reporting requirements and practices from facilities to the system level. It also discusses factors arising from regulations, including classifications and standardization.

**Figure 1.**
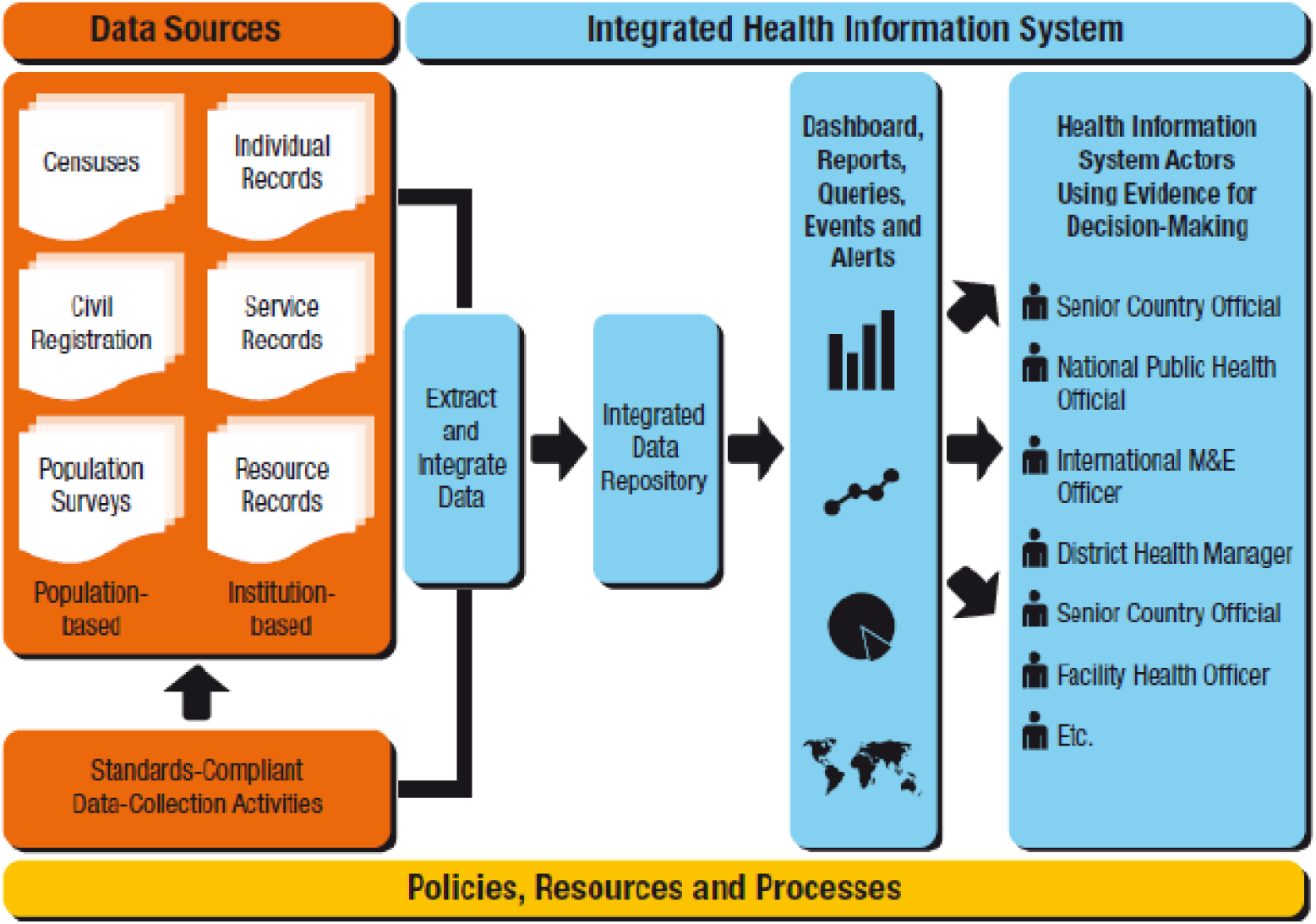
Health Metrics Network’s framework for health information system. Source: World Health Organization

**Figure 2.**
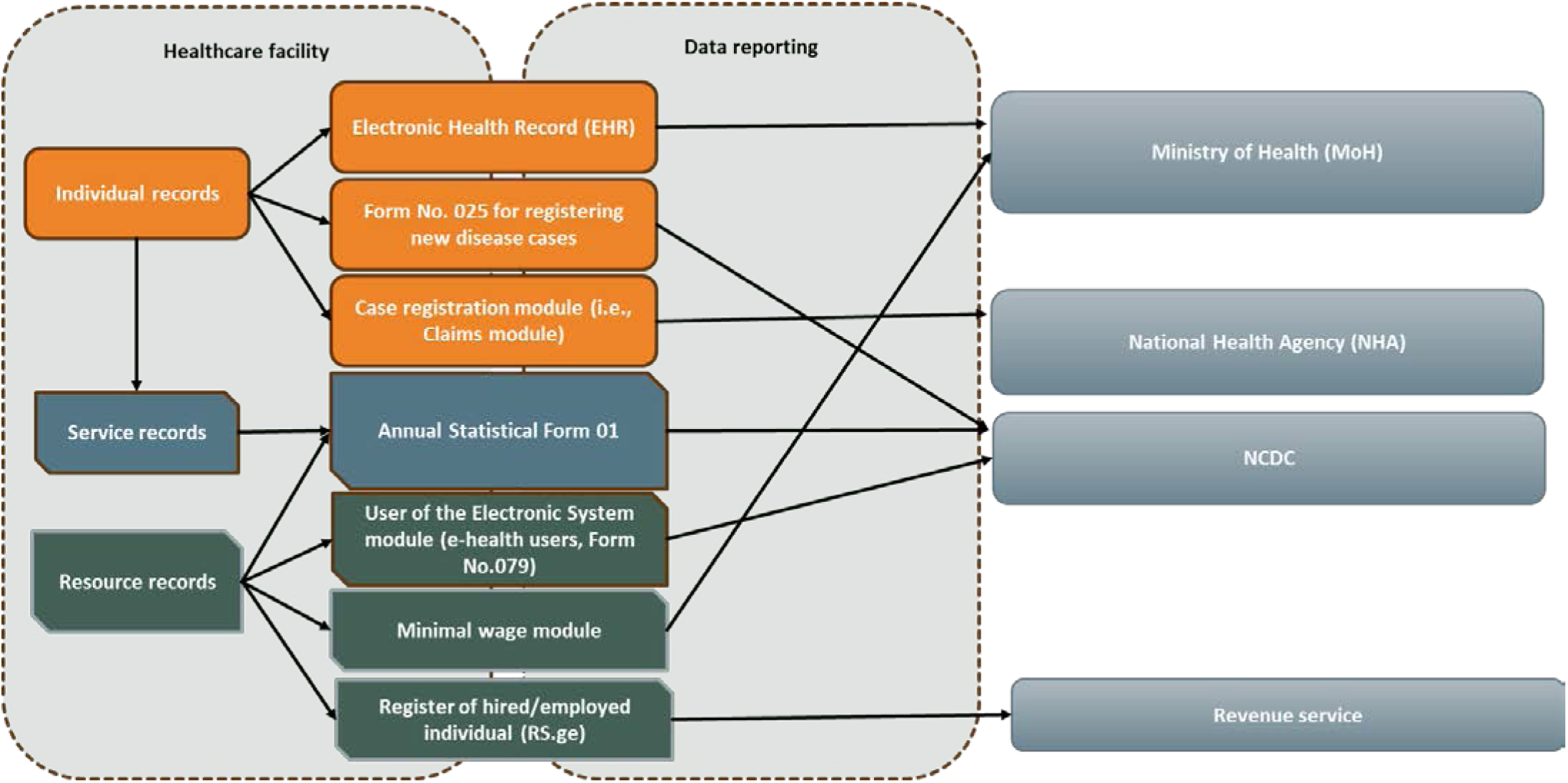
Data reporting from the facility level to the system level. Source: compiled by authors

Rehabilitation service providers must submit data through seven specific modules/forms. Figure 2 illustrates the reporting forms/modules and processes from the facility to the system level, showing the forms and corresponding entities responsible for data receipt and management.

Facility-level reporting forms include:

- Electronic Health Record (EHR), which is submitted to MoH
- Form 025, used to register new outpatient cases, submitted to the NCDC
- Case registration module (i.e., Claims module), submitted to the NHA
- Annual Statistical Form 01, submitted to the NCDC
- User of the Electronic System module (e-health users, Form No.079) submitted to the NCDC
- Minimal wage module, submitted to MoH
- Register of hired/employed individuals (RS.ge), submitted to the revenue service of the Ministry of Finance for income tax management purposes.

Each form falls into one of three categories—individual records, facility records, and resource records—and each faces unique challenges related to data volume, reporting standards, and analytical complexity, as described below.

### Individual-patient records

The practice of data collection on a facility level is variable in terms of the type and volume of information collected and then transferred/reported through the system. Namely, significant discrepancies exist among facilities contracted under the UHCP. Some UHCP providers record and report complete information about all rehabilitation episodes in EHR, from the initial visit to the final, while others record completed data for a patient but report only data for the initial visit. As one respondent noted:

> *“When the first consultation is conducted, it [the data] is sent to the EHR, but the final record is not submitted [through this system]” (Respondent 2)*.

Conversely, another respondent explained:

> *“When a beneficiary’s episode is concluded, the EHR requires reporting within fourteen calendar days from the end of the episode. This includes all visits and procedures performed during that time [throughout rehabilitation]” (Respondent 9)*.

In general, recording requirements for facilities outside UHCP are governed by similar rules (the same MoH decrees). Nonetheless, reporting requirements vary and depend on the form and entity that requires such reports. Namely, all NCDC-required forms are supplied by all facilities in the country delivering rehabilitation service – a universal mandate. However, forms required by UHCP/NHA and MoH (such as claims module, EHR and minimal wage module) are not compulsory for the facilities operating outside of UHCP. This also explains the variable practice in data collection/recording on a facility level and the eventual reporting observed among UHCP and non-UHCP facilities (see Figure 3).

**Figure 3.**
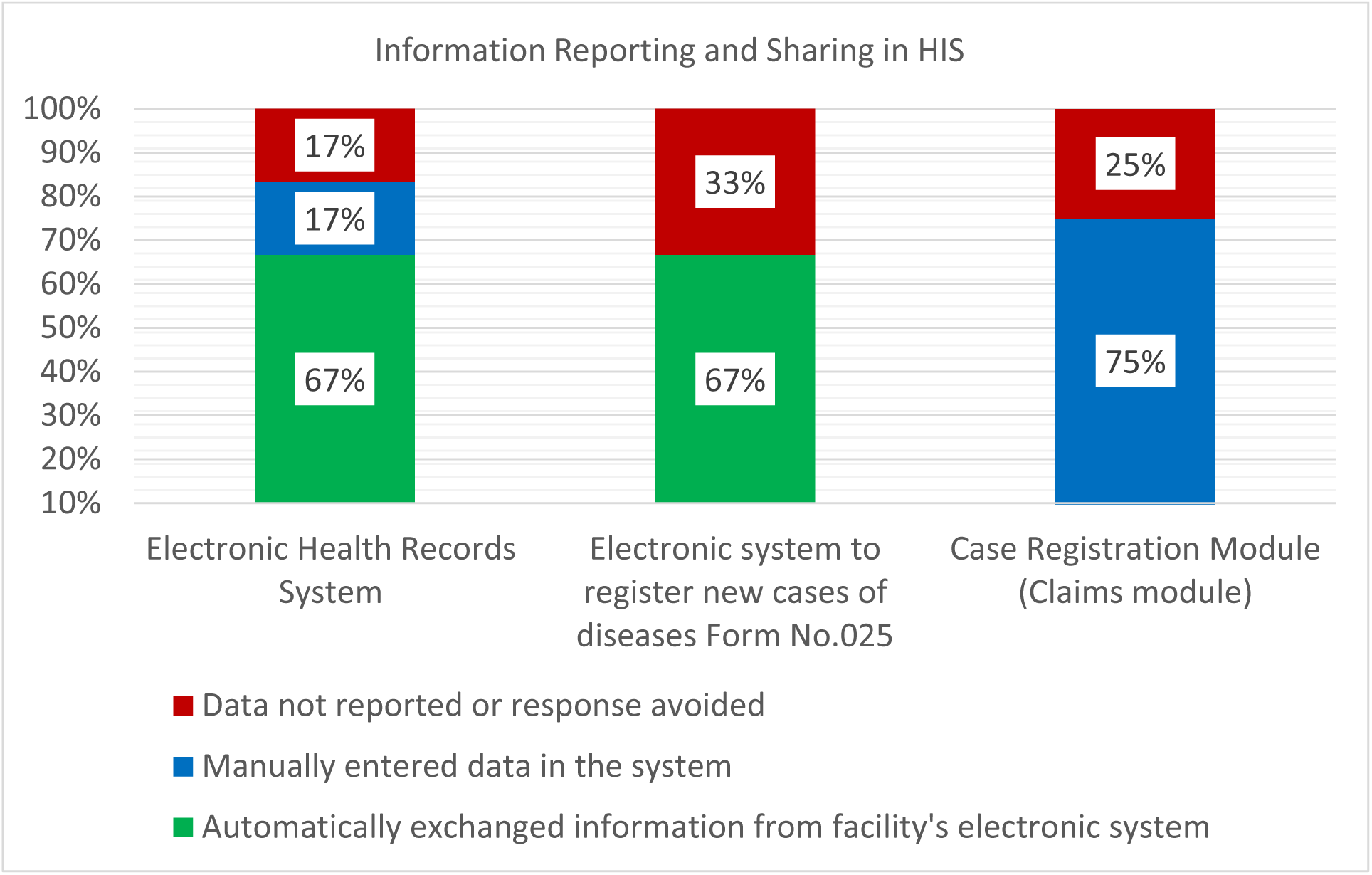
Information reporting and sharing practices in HIS

Figure 3 presents data derived from a study of six facilities. In the Case Registration Module, data reporting is limited to UHCP providers, with four UHCP facilities included in the analysis.

Respondents identified weakly formulated regulations and lack of guidance from state entities as key reasons for data collection/ recording and reporting inconsistencies.

> *“When the rehabilitation program started, there were no official guidelines or healthcare protocols, so we were left to manage the process ourselves. There was only one order explaining how to qualify as providers and meet the program’s criteria” (Respondent 1)*

In addition, inconsistencies were noted in the reporting of cost-of-service data. Some facilities report this data in the EHR financial module, while others omit financial details when supplying medical and patient data in the EHR. Also, those that report cost data in the EHR classify the cost data differently due to a lack of clear methodological guidance, which limits the comparability and, therefore, the utility of collected financial information. As another respondent noted:

> *“These details have not been clarified with the Ministry, and we still lack contact with anyone at the MoH regarding these issues. As a result, we recorded what we could in the EHR, despite the limitations” (Respondent 9)*.

Additional shortcomings were uncovered in data recording (and consequently in reporting) standards. Namely, the system lacks standards/classification for measuring the degree of functional impairment of the patient. Although all facilities use the International Classification of Diseases, 10th Revision (ICD-10), no unified classification system exists for recording functional impairments or the type of rehabilitation interventions delivered to the patient. Facilities participating in state programs must use the *Nomesco Classification of Surgical Procedures* (NCSP) to record interventions, but others outside of UHCP do not use any classification system for intervention coding, which undermines data comparability across the facilities and impedes the possibility of monitoring the current levels of service delivery or planning the future needs. NCSP classification was introduced in Georgia almost two decades ago and has not been routinely updated since. Therefore, the international codes are missing for some rehabilitative interventions, and MoH introduced non-standardized “artificial” codes in the case registration module instead of internationally recognized ones. One respondent remarked,

> *“The case is reported in the system [case registration module] when the patient comes in. The procedures are recorded using ‘artificial codes’ (designed by the Ministry of Health) along with the diagnosis using ICD-10 codes” (Respondent 10)*.

Furthermore, practices of assessing the functional status of a patient are also inconsistent. Georgia has not adopted and institutionalized international classifications, such as the *International Classification of Functioning, Disability, and Health* (ICF) (12), which constrain the ability of the state to produce routine statistics and monitor the populations’ rehabilitative needs. Although the state mandates the use of the *Functional Independence Measure* (FIM) exclusively for UHCP beneficiaries (25). some UHCP providers also use additional assessment tools for internal purposes. As one respondent explained:

> *The assessment is conducted using the patient’s Functional Independence Measure (FIM). We use specific FIM scores to evaluate the patient at the start of rehabilitation. Afterward [after completing a course of rehabilitation], we submit a patient summary, including final assessment score, to the Ministry of Health” (Respondent 10)*.

In contrast, facilities delivering services outside UHCP rely on non-standardized and non-uniform functional assessment tools of their own choice. For instance, some non-UHCP providers use their own adapted assessment instruments. One respondent described their facility’s custom system:

> *“We have implemented our own clinical score system to track changes in a patient’s symptoms over time. This system allows us to assess the progression of their condition—for example, whether pain has subsided, or mobility has improved. We document changes such as ‘he couldn’t, and now he can,’ to capture these dynamics” (Respondent 16)*.

The use of variable assessment instruments (or the lack thereof) challenges comparability between UHCP and non-UHCP facilities, limiting the State’s ability to monitor rehabilitation outcomes on a national level.

The next challenge arises from the format of the reporting requirements placed on providers. For example, for UHCP beneficiaries, FIM data is being reported in PDF format to NHA instead of having dedicated data fields in the system for initial assessment and separately for the outcome of rehabilitative interventions, complicating the retrieval and analysis of beneficiaries’ functional improvement outcomes on a program level. As noted by respondents,

> *“Patient summary (form 100) includes the initial Functional Independence Measure (FIM) score established at the first visit and last visit dates, the [rehabilitation] interventions performed, and the final FIM score [to reveal the outcome of rehabilitation]” (Respondent 1)*.

Another challenge within the system is the duplication of data reporting across different reporting modules, which places a significant compliance burden on facilities. For instance, the EHR system contains 126 variables, while Form 025 requires 25 variables, of which 18 overlap with the EHR system. This redundancy leads to approximately 80% of information duplication between Form 025 and the EHR system (see Figure 4). This redundancy originates from outdated reporting requirements and a lack of integration across systems. Despite capturing the same information in the EHR, facilities must re-enter data into Form 025 to comply with state regulations. This duplication strains facility resources and staff time, redirecting focus from patient care to administrative tasks.

**Figure 4.**
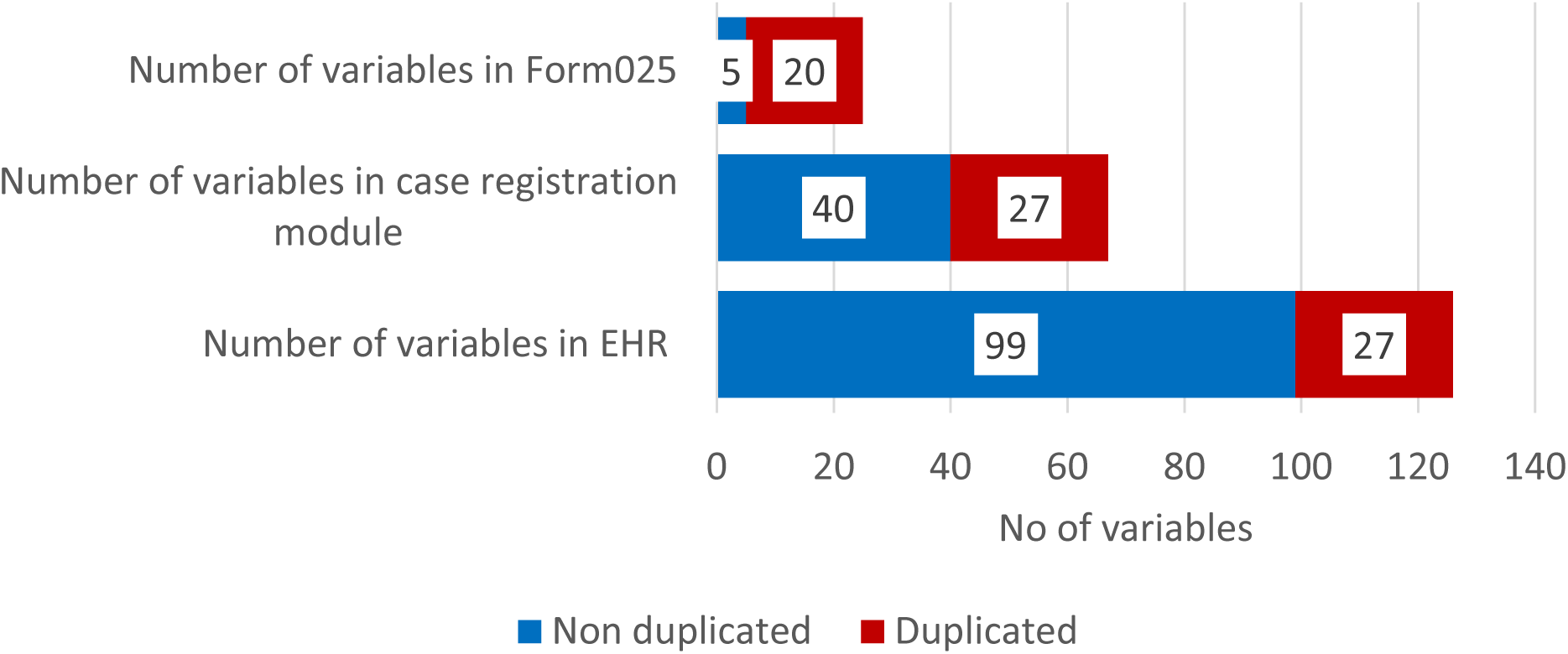
Data duplication in key health information modules

## Facility records and Human resources data

Data reporting inconsistencies exist not only in individual records but also emerge in facility and human resources reporting forms. As illustrated in Figure 5, reporting practices across facilities vary widely highlighting the fragmented nature of the HIS. The data in Figure 5, derived from a study of six facilities. Data reporting in the minimum hourly wage module is limited to UHCP providers, with four UHCP facilities included in the analysis.

**Figure 5.**
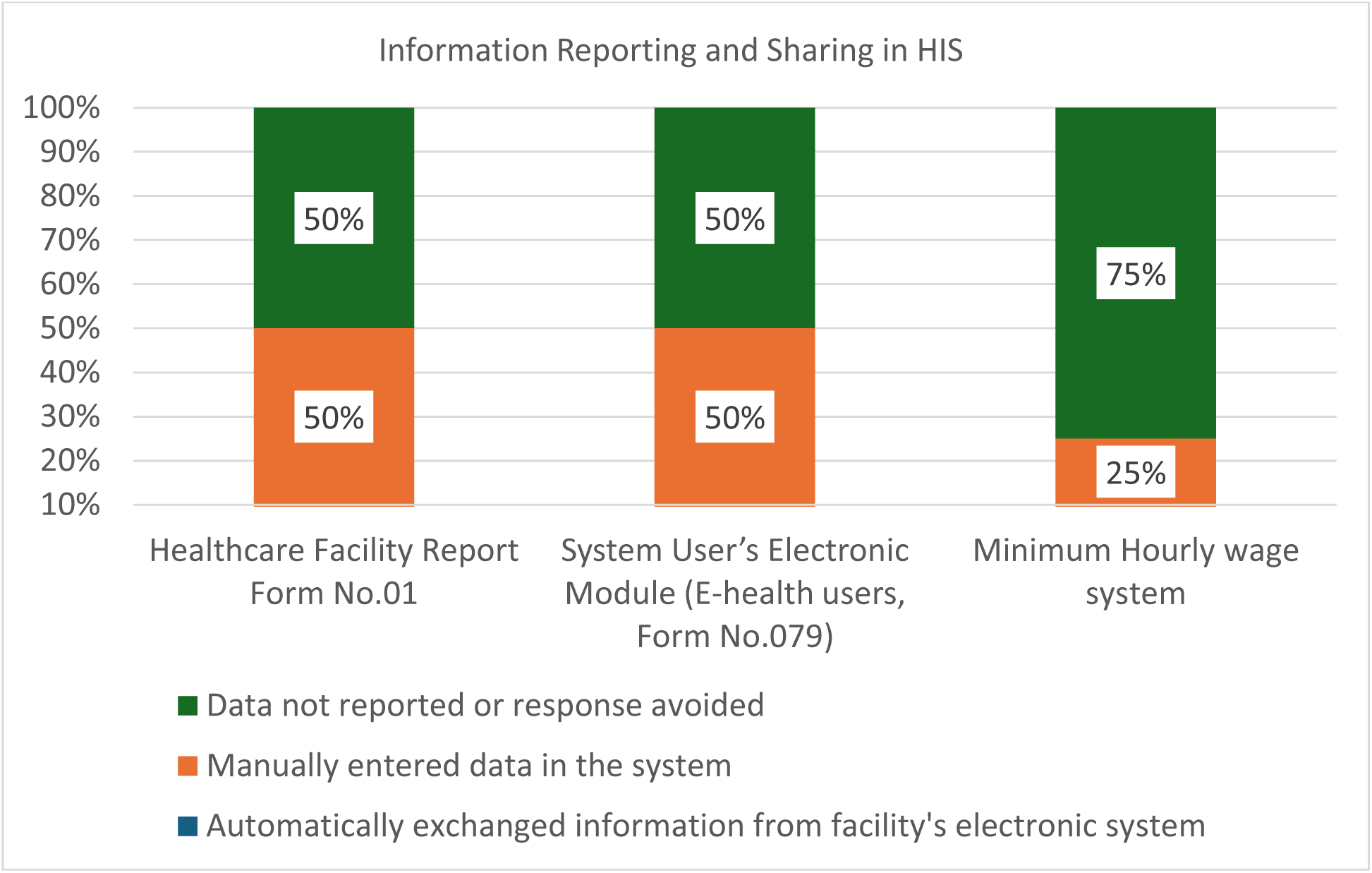
Information reporting and sharing practices in HIS

Facility records are collected annually through the Healthcare Facility Report (Form No. 01), which includes data on equipment, staffing, and service volume. For rehabilitation services, the form records the number of employees and activities in departments such as physiotherapy, medical physical culture, and reflexotherapy. It also tracks the total number of patients who completed a rehabilitation course and the number of interventions performed.

Human resource data is collected through three different modules. First, The Electronic System Module (e-health/ Form No 079), introduced by the Ministry of Health during the COVID-19 pandemic, provides real-time data on healthcare personnel. This system tracks employed personnel through unique national identification numbers (IDs), allowing for up-to-date analysis of active doctors and nurses employed by facilities.

Second, The Minimum Wage Module monitors employment standards by recording hours worked and wages earned, ensuring compliance with MoH minimum wage regulations and promoting fair labor practices. However, the regulations are only targeted at clinics contracted under the UHCP.

Additionally, according to a directive from the Ministry of Finance in 2010, employers are required to submit monthly reports detailing employee information, their salaries, income tax deduction and any changes in employment status updated within five days of occurrence. This electronic register was strengthened on February 1, 2021, to ensure more consistent reporting practices (26,27).

Despite these multiple reporting modules, data duplication and inconsistencies persist across human resource modules. For example, in the Minimum Wage System, 9 out of 24 variables are duplicated, while the Electronic System Module (e-health/ Form No 079) has 9 duplicated variables out of 27, and the RS.ge system shows 5 out of 7 variables are duplicated. These issues highlight uncoordinated approach of the parts of the government imposing double reporting burden on the providers and ongoing challenges in ensuring accurate and consistent data across the various reporting systems/modules (see Figure 6).

**Figure 6.**
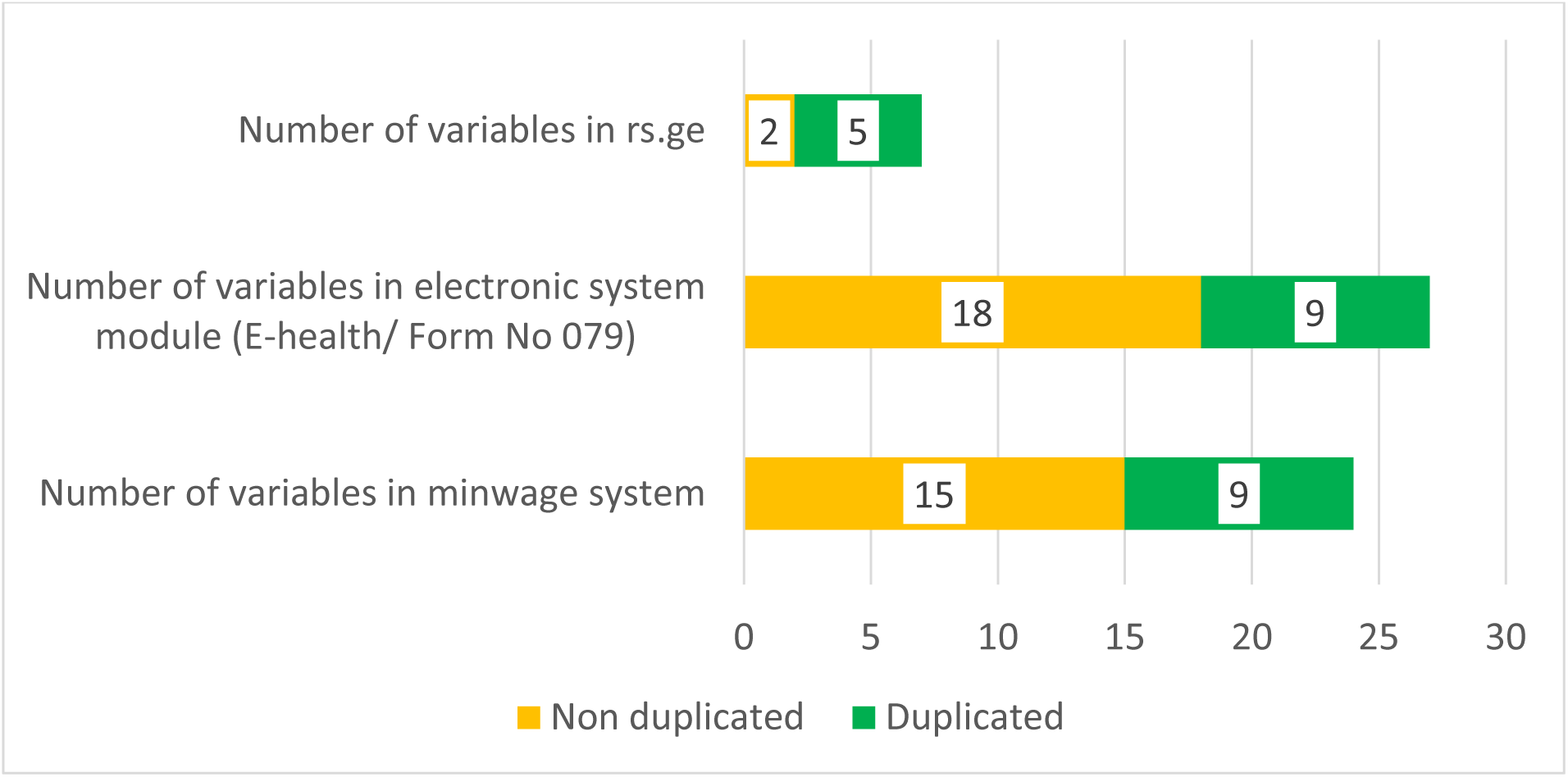
Comparison of data duplication in HR modules.

## Discussion

Data related to disability and rehabilitation are often missing from HIS, particularly in LMICs, where health information systems are generally weak (28). Our study shows that Georgia is not an exception and raises the critical question: Are the current HIS sufficient to support nationally established indicators? What factors impede the system’s functionality in this regard? What needs to happen to improve it?

Evidence highlights common barriers to effective HIS include systemic instability, unclear roles, fragmented political responsibilities, and complicated decision-making processes. These issues are well-documented not only in LMICs but also in European contexts (29). Our study shows that the absence of a robust regulatory framework in Georgia has led to inconsistent data collection, poor classification, and a lack of data quality and standardized reporting practices across facilities.

An essential step to improving data quality is using electronic data collection tools, such as HER (30). While Georgia has made progress with EHR implementation, significant challenges remain. One major issue is the variability in the type and volume of information collected and reported. This underscores the need for Minimum Data Sets (MDSs) for both UHCP and non-UHCP providers, as the absence of MDSs hampers data completeness and comparability both nationally and internationally (31,32).

Our study also identified the absence of a unified, nationally adopted classification system for measuring functional impairment and recording rehabilitative interventions. Internationally, the ICF is widely accepted for categorizing functioning and disability. Combining the ICF with the International Classification of Diseases, 11th Revision (ICD-11) for disease recording and the International Classification of Health Interventions (ICHI) for coding interventions is recommended (33,34). However, Georgia has not yet implemented the ICF or ICHI. Currently, functional assessments for UHCP beneficiaries primarily use the FIM, whereas no standardized approach exists for other cases. Some facilities use NCSP codes to record rehabilitation interventions, but this practice is inconsistent. A phased approach is recommended to improve data recording practices. Initially, mandating functional outcome measures across all rehabilitation facilities and standardizing the use of NCSP codes would establish a foundation for consistent and comparable data collection. In the subsequent phase, adopting the ICF and ICHI would enhance alignment with international standards, thereby improving the quality of health data and enabling better cross-national comparisons.

It is crucial to emphasize that adopting international classification systems without establishing standardized recording and reporting formats will undermine the utility of the data collected. Our findings reveal that the absence of structured digital reporting formats poses significant data extraction, analysis, and utilization challenges. For example, submitting FIM and financial data to the National Health Agency in PDF format impedes data analysis, hindering the evaluation of rehabilitation outcomes, program effectiveness, and economic performance.

Another critical challenge is the lack of interoperability between various reporting systems, which creates administrative burdens for healthcare facilities. Many facilities are required to report duplicate data across multiple modules, leading to inefficiencies, particularly for those with lower service volumes. Addressing these challenges necessitates the establishment of standardized protocols not only for data collection but also for system interoperability. Adopting internationally recognized healthcare interoperability standards such as Health Level 7 (HL7), Fast Healthcare Interoperability Resources (FHIR), Digital Imaging and Communications in Medicine (DICOM), Clinical Document Architecture (CDA), and SNOMED-CT can facilitate seamless communication between systems (35). Addressing these challenges necessitates not only the adoption of internationally recognized standards but also concerted efforts to improve systems integration, resolve regulatory conflicts, and eliminate data silos. By focusing on these critical areas, Georgia can make substantial progress in creating a more efficient and effective HIS for rehabilitation service delivery.

This study has certain limitations. The reliance on in-depth interviews resulted in a small sample size. The study covered 67% of UHCP rehabilitation providers (two in the capital city and two in the regions) with additional input from two facilities operating outside the UHCP, limiting the findings’ generalizability. Additionally, the purposive sampling approach may have introduced selection bias, as facilities with more advanced data management practices might be more likely to participate in the UHCP or possess better data capabilities. While the qualitative nature of the study provided rich insights, it also limited the ability to draw statistically generalizable conclusions. Despite these limitations, the use of triangulation and regular coding reviews strengthened the reliability of the findings. To further address these limitations, validation workshops were conducted with diverse stakeholders, including policymakers and rehabilitation providers, ensuring the findings and recommendations were relevant and robust and reflected general situation in the country.

## Conclusion

In conclusion, while Georgia has made notable progress in integrating rehabilitation services into the UHCP, its HIS faces significant challenges. Critical gaps, such as non-standardized data collection, inconsistent data standards, and poor interoperability, limit the ability to strategically plan, evaluate, and expand rehabilitation services. Developing a standardized data collection framework focused on functional outcomes, patient-centered measures, and the comprehensive use of electronic systems is essential to address these issues. Strengthening the HIS through coordinated efforts and a clear regulatory framework will enhance service delivery and provide valuable insights for LMICs striving to optimize rehabilitation services. Further research is needed to explore how rehabilitation data can be fully integrated into HIS to improve patient outcomes and support evidence-based policymaking.

## Data Availability

All data produced in this study are available from the authors upon reasonable request.

## Notes

### Competing Interest Statement

The authors have declared no competing interest.

### Funding Statement

This research was supported by Results for Development (R4D), whose grant funded the time and resources necessary for data collection and analysis.

### Author Declarations

Ethical approval was obtained from the National Bioethics Committee of Georgia (Ethical approval (IRB # 2023-074)) housed at the National Center for Disease Control and Public Health (NCDC) before the commencement of the fieldwork.

